# Multidimensional evaluation of the early emergence of executive function and development in Bangladeshi children using nutritional and psychosocial intervention: A randomized controlled trial protocol

**DOI:** 10.1101/2023.12.16.23300075

**Authors:** T. Shama, J.M. O’Sullivan, N. Rahman, S.H. Kakon, F. Tofail, M.I. Hossain, M. Zeilani, R. Haque, P. Gluckman, T. Forrester, C.A. Nelson

**Author notes:** Co-corresponding authors: Justin O’Sullivan; Rashidul Haque,; Terrence Forrester,; Chuck Nelson,.

## Abstract

**Introduction:** Reversing malnutrition-induced impairment of cognition and emotional regulation is a critical global gap. We hypothesize that brain-targeted micronutrient supplemented nutritional rehabilitation in children with moderate acute malnutrition, followed by 2 years micronutrient supplementation will impact on the cognition and emotion regulation of these children.

**Methods:** The primary outcome of this prospective, randomized controlled trial is to study the development of executive functions (EFs) and emotion regulation (ER) in this cohort.

Moderate acute malnourished (MAM; WLZ/WHZ <-2 and ≥-3 z-score, and/or 11.5 cm ≤ MUAC < 12.5cm; n=140)children aged around one year (11m-13m) in Mirpur, Dhaka, Bangladesh will be randomized (1:1) to receive either locally produced Ready to Use Supplementary Food (RUSF) or Enhanced Ready to Use Supplementary Food (E-RUSF) until anthropometric recovery (WLZ/WHZ > -1SD), or for 3 months after enrollment (whichever is earlier). The randomized MAMs groups will be given either Small Quantity Lipid Based Nutrient Supplement (SQLNS) or Enhanced Small Quantity Lipid Based Nutrient Supplement (E-SQLNS), respectively until the end of the 2-year follow up period. Standard psychosocial stimulation will be provided to the MAMs intervention groups. Biological samples will be collected, anthropometric and neurocognitive assessments will be performed at 2 (22m-26m) and 3 (34m-38m) years of age.

Two control groups will be recruited: 1), non-malnourished one-year (11m-13m) old children (WLZ/WHZ score>-1SD; n=70); and 2) three –year (34m-38m) old children (n=70) with untreated MAM (WHZ <-2 and ≥-3 z-score, and/or 11.5≤MUAC<12.5 cm). The 3-year-old MAM reference group will be assessed once and provided with 2 months of nutritional rehabilitation support (RUSF Nutriset’s Plumpy’Sup^TM^).

**Ethics and dissemination:** The study protocol has been reviewed and approved by all the relevant ethical review boards at each research site (icddr,b Bangladesh; University of the West Indies, Kingston Jamaica; and University of Auckland, New Zealand). The results will be disseminated through peer-reviewed publications and national and international scientific conferences.

Protocol version 1 (1/11/2022)

**Trial registration number:** NCT05629624. Registered on November 29, 2022.

## Introduction

### Background and rationale

Nutrition is essential at all stages of life, beginning before birth and continuing throughout childhood. During these periods, critical developmental windows facilitate healthy brain development, including their behavioral correlates (particularly cognitive, motor, and socio-emotional skills [1]. In low- and middle-income countries (LMICs), 45% deaths are due to undernutrition, where around 2 billion survivors suffer long-term cognitive and behavioral consequences [2]. While less than half of all children under-five live in LMICs, nearly 66% of all stunted and 75% of all wasted children live in this region. The highest global prevalence of stunting is found in Asia, with 53% of all stunted children, and 70% of all wasted children located in Southern Asia [3, 4], making this region a high priority for intervention. Since 2000, stunting has steadily declined but a high prevalence of wasting persists hindering the achievement of Sustainable Development Goal 2: zero hunger targets by 2030 [5, 6], an important marker for development in this region.

Bangladesh, situated in the Indian subcontinent bordering the Bay of Bengal, is amongst the most densely populated countries globally [7]. Malnutrition is a significant health issue, with approximately one-third of preschool-age Bangladeshi children being stunted (low height-for-age), ≥ one-fifth are underweight (low weight-for-age) and approximately one-tenth are wasted (low weight-for-height) [8–10]. In Bangladesh, approximately 1.8 million children under 5 years of age suffer from Moderate acute malnutrition (MAM) [11–13], diagnosed as weight-for-height (WHZ) or weight for length (WLZ) z-scores between <-2 and ≥-3 of WHO child growth standards and/or mid upper arm circumference (MUAC) <12.5 and ≥11.5 cm [14, 15].

Malnourished children have multiple micronutrient and macronutrient deficiencies and exhibit an abnormal gut microbiota [16], which can disrupt the bidirectional neural and immune interactions between the gut and brain by affecting signal molecules produced by the microbiota (e.g. short-chain fatty acids, and neurotransmitters) [17]. Nutritionally wasted children exhibit brain atrophy on MRI, and while re-feeding reverses the shrinkage of the brain, significant brain microstructural and functional deficits persist [18]. The anatomic reconstitution of the brain with feeds designed principally for rapid physical catch-up growth might result in atypical cognitive and emotional performance. Correcting cognitive and emotion regulation outcomes after malnutrition requires providing the appropriate nutrients in amounts to support rapid catch-up growth of both the body and brain.

The human brain develops from prenatal life through early childhood, with a crucial developmental window between birth and age 3 years [19]. Malnutrition, which includes nutrient deficiencies, and gut microbiome dysbiosis can induce structural and functional abnormalities of the brain that lead to lifelong neuropsychological sequelae [20, 21]. Improving brain recovery in malnourished children might require augmented feeds with key nutrients that have targeted functionality in the brain, particularly during periods of rapid development.

## Methods

### Study setting

This study is a community based randomized controlled clinical trial, conducted in the Mirpur area of Dhaka, Bangladesh. Mirpur is divided into five zones, with a population of approximately half a million people. The area is densely populated, with > 38,000 people living in each square kilometer [22], in contrast to Tokyo which has 7,330.6 persons per square kilometer. The Dhaka cohort represents a typical urban-poor population exposed to poor living and sanitary conditions, environmental hazards, toxins, and poor access to nutrition.

All clinical activities will be conducted at the Mirpur field clinic, which is located in ward 5, in close proximity to the participants. This clinic has been a research site for nearly 30 years.

This study protocol was registered on clinicaltrials.gov on November 29, 2022 (study ID number: NCT05629624) and was produced in compliance with the Standard Protocol Items: Recommendations for Interventional Trials (SPIRIT). See Fig 1 summarizing the schedules of enrollment, interventions and assessments of the study.

**Fig. 1.**
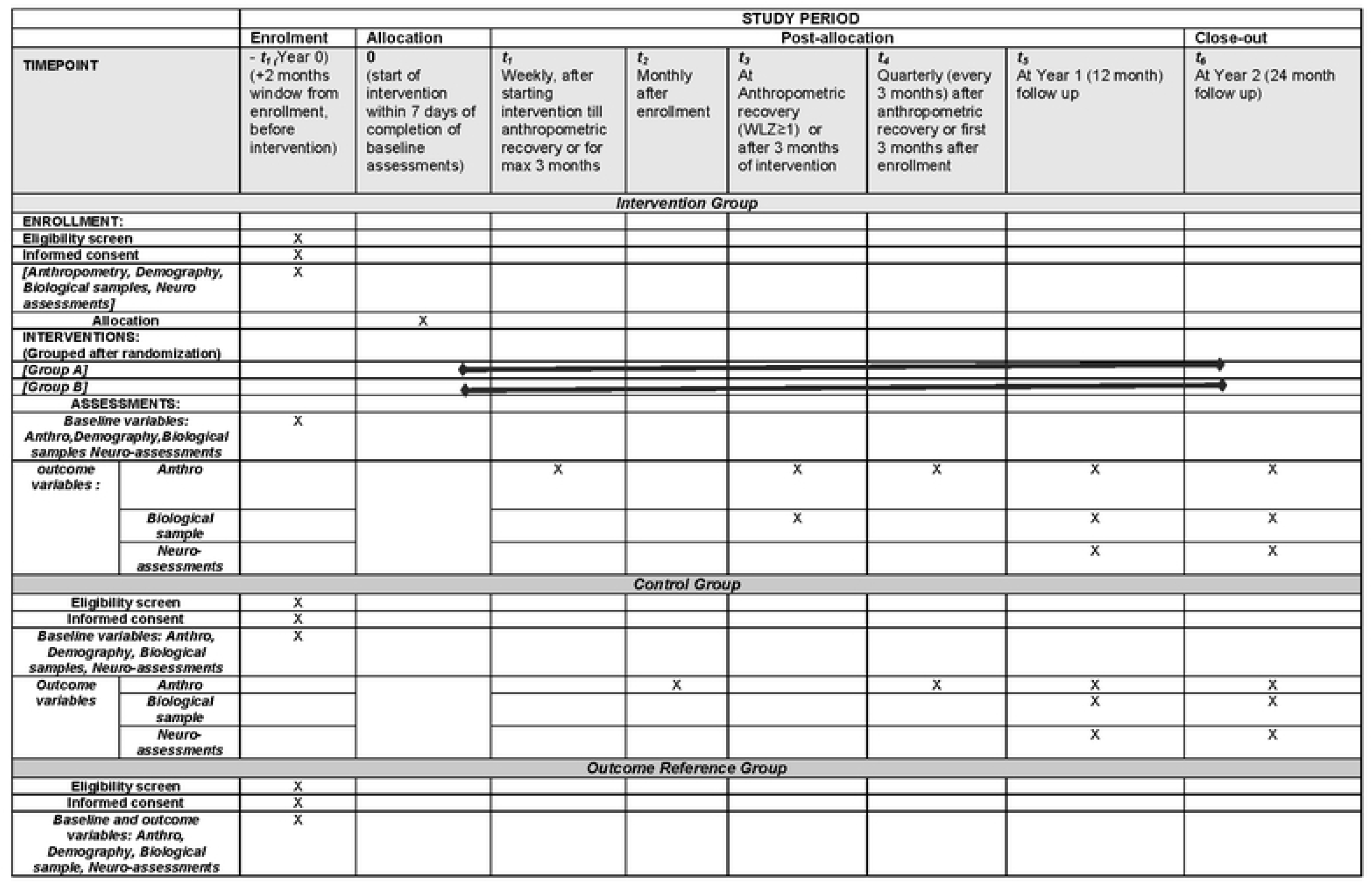
SPIRIT schedule. Schedules of enrollment, interventions and assessments

### Objectives and trial design

We propose an enhanced nutritional intervention to investigate the impact of enriching the diet with micronutrients and prebiotics during recovery from MAMs on executive functions (EFs) and emotion regulation (ER). This intervention will be combined with psycosocial stimulation. The study cohort will be recruited within the Mirpur locality in Dhaka city, Dhaka North City Corporation (DNCC) wards 2, 3 and 5.

Our primary outcome is to study the development of EFs and ER in this cohort. The secondary outcomes are to assess whether nutritional interventions and psychosocial stimulation improve executive dysfunction (EDF) and emotion dysregulation (EDR) upon recovery from childhood moderate acute malnutrition. We will identify features of microbiome assembly, metabolomics, and child genetics that respond to nutritional intervention and contribute to executive and emotion regulation/dysregulation in healthy Bangladeshi children. The findings from this study will identify the effects of nutritional intervention and psychosocial stimulation on EF and ER in malnourished children, thereby offering valuable insights for scalable approaches to improve these aspects of child development.

### Recruitment

Enrollment was initiated on February 7, 2022 and will continue until February 2024.

Study surveillance workers (SWs) conducted a door-to-door census (∼ 100,000 households) in Mirpur DNCC wards ward 2, 3 and 5 (see the area map in Fig 2) between January and December 2022. The primary focus of the census was to compile a list of babies aged 11 to 13 months and 34 to 38 months. Verbal consent was obtained to participate in the census. The census identified 5,736 children aged between 11 to 13 months and 2,314 children aged between 34 to 38 months. During the census, if the guardian verbally consented to the study procedure, and the babies met the inclusion and exclusion criteria of the study (Table 1), the SWs proceeded to measure the mid-upper arm circumference (MUAC) of the child. We are inviting mothers of the babies who are within the expected MUAC range to visit the study clinic for further assessment and enrollment.

**Fig. 2.**
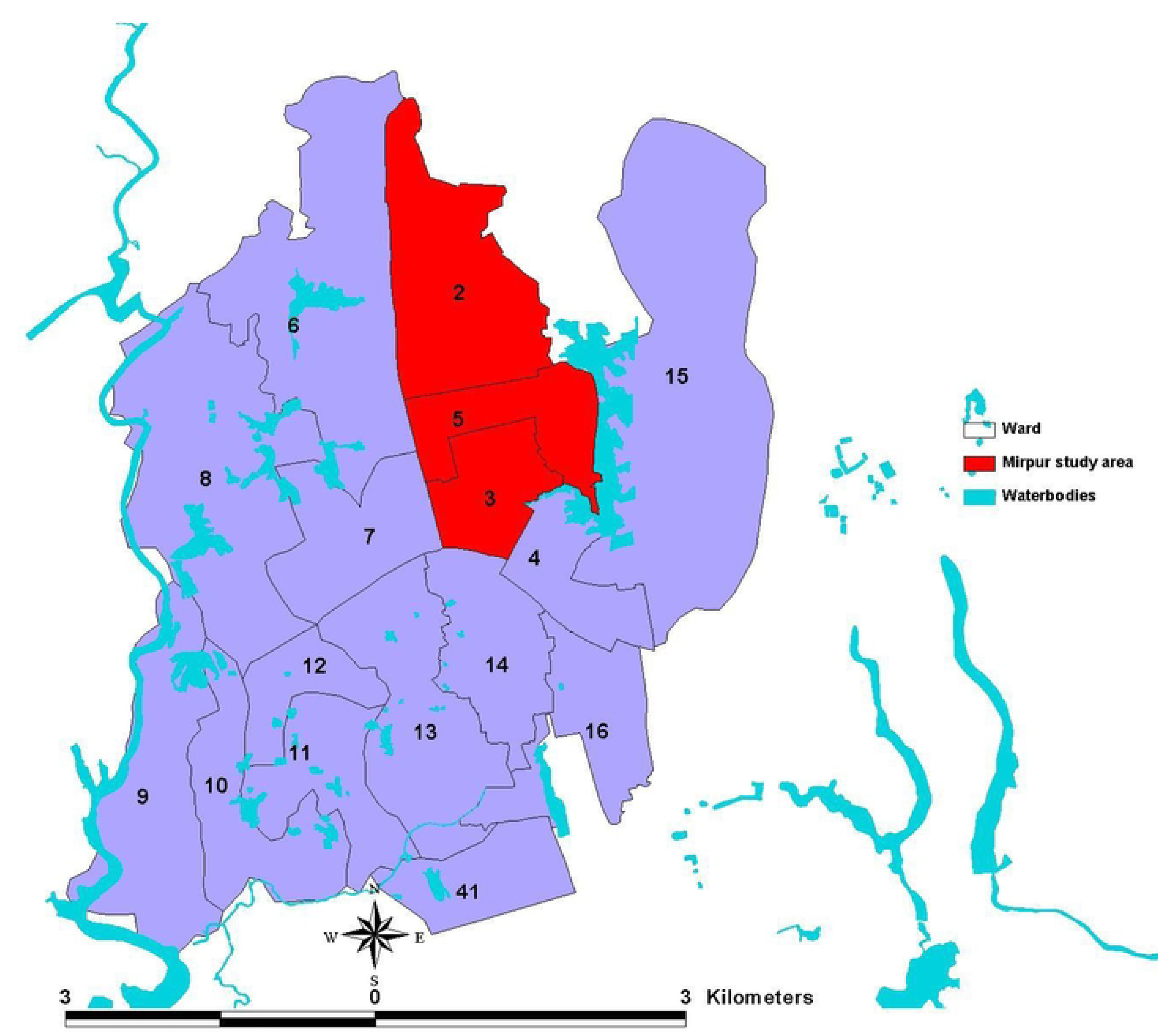
Study area at Mirpur includes Ward 2, Ward 3 and Ward 5.

**Table 1.** Eligibility criteria.

| **Intervention group**<br>**1 year MAM (n=140)** | **Control group**<br>**1 year well-nourished (n=70)** | **Outcome reference group**<br>**3 years MAM (n=70)** |
| --- | --- | --- |
| *Inclusion Criteria:* |  |  |
| Child aged between 1year ± 1m (11m - 13m).<br>Child's WLZ/WHZ score is $<-2$ and $\geq-3$, and/or MUAC $<12.5$ and $\geq 11.5$ cm. Child is free from any acute illness.<br>Mother is willing to sign informed consent form. | Child's aged between 1 year ± 1m (11m - 13m).<br>Child's WLZ/WHZ score is $> -1$ SD, Child is free from any acute illness.<br>Mother is willing to sign informed consent form. | Child aged between 3 year ± 2m (34m – 38m).<br>Child's WLZ/WHZ score is $<-2$ and $\geq-3$, and/or MUAC $<12.5$ and $\geq 11.5$ cm. Child is free from any acute illness.<br>Mother is willing to sign informed consent form. |
| *Exclusion Criteria:* |  |  |
| Child aged $> 13$ months or $< 11$ months.<br>Child has any congenital anomaly.<br>Mother is unwilling to sign informed consent form.<br>Mother unwilling to bring child to the clinic for assessments or to use intervention feeds on child. | Child aged $> 13$ months or $< 11$ months.<br>Child has any congenital anomaly.<br>Mother is unwilling to sign informed consent form.<br>Mother unwilling to bring child to the clinic for assessments | Child aged $> 38$ months or $< 34$ months.<br>Child has any congenital anomaly.<br>Mother is unwilling to sign informed consent form.<br>Mother unwilling to use the rehabilitation feed on child or the small quantity supplement. |

Final screening for eligibility and study consent will occur at the Mirpur study clinic. Because the primary study population is incapable of assenting, due to age 1year ± 1m and 3 years ± 2m, parental permission will be obtained for study participants. The consenting process will be done by a study physician. The consenting process will be tailored to each mother’s literacy level and will involve reviewing the inclusion and exclusion criteria. Comprehension of the study will be assessed using scripted points and open-ended questions. After obtaining consent, the clinical screening team will proceed with anthropometric screening of the child and mother.

Following consent, the clinical screening team will complete a screening form (Supplementary materials 1), capturing the date of visit, sex, date of birth (DOB), weight (in kg), length/ height (in cm), head circumference (in cm), and MUAC (in cm) measurements of the child. The team will calculate the WLZ/WHZ Z-score for the child using the WHO anthropometric calculator. The child’s age will be validated using the EPI vaccination card.

Final enrollment eligibility is based on matching the Z-score and other physical examination findings with the eligibility criteria (Table 1).

### Control Groups

Two control groups will be recruited. The first control group will consist of non-malnourished children who are recruited at the same age (11 months to 13 months old) as the MAM intervention group children.

The second control group (i.e. outcome reference group) will be a MAM reference group consisting of children who are 3 years (34 months to 38 months old) who present with previously untreated MAM. See Fig 3 demonstrating the flow of recruitment process of the study participants.

**Fig. 3.**
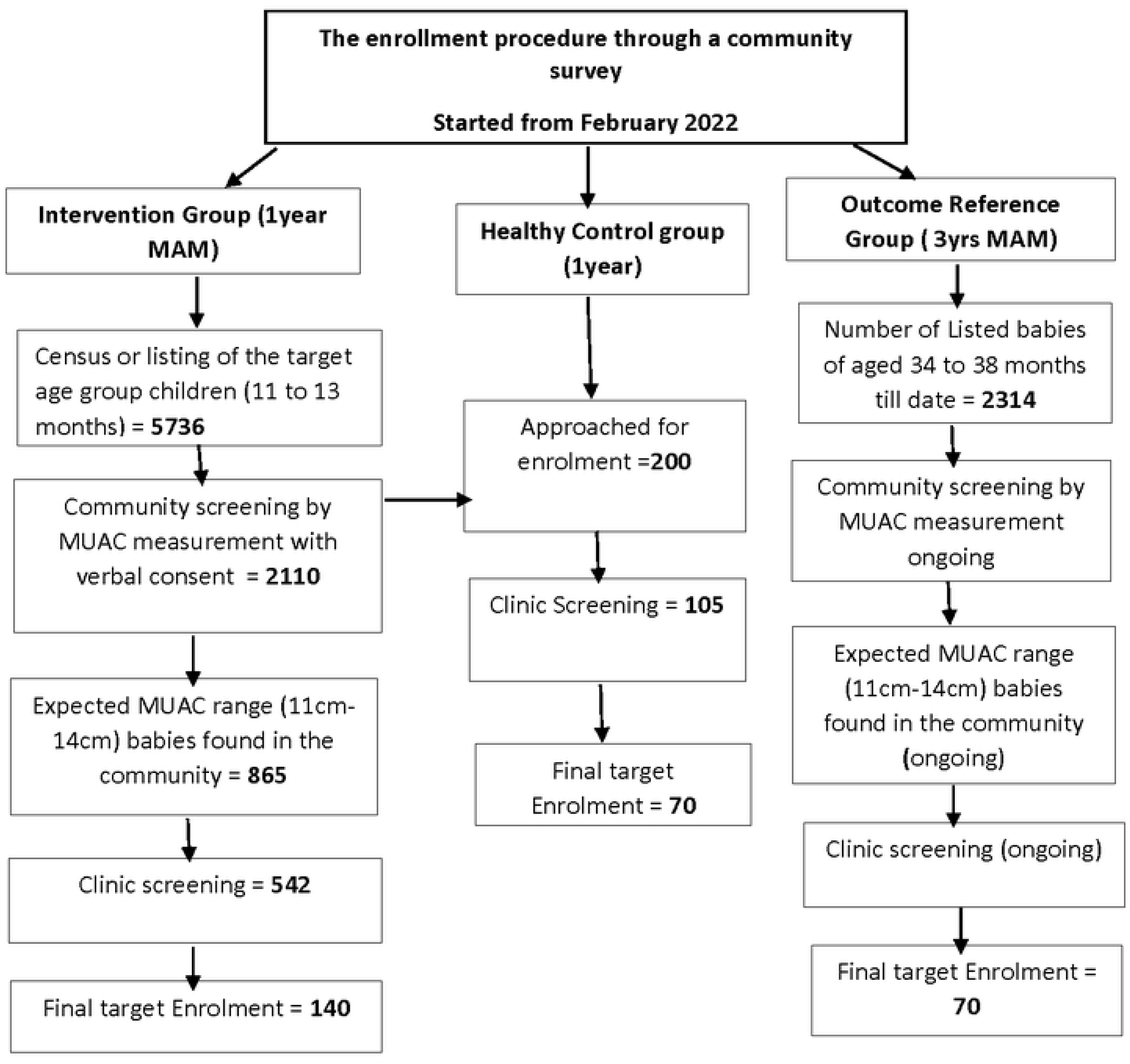
Flow chart of the study settings and participants

### Sample size

Power for comparisons between well-nourished (WLZ/WHZ > -1 SD) and wasted children (-3≤ WLZ/WHZ <-2 z-score, and/or 11.5≤ MUAC <12.5 cm) at 1 and 3 years was estimated using the Student’s t-test and Structural Equation Modeling (SEM). For the t-test, the proposed medium level difference between the two groups (i.e. 2 standard deviation) requires a minimum of 51 participants per group for power equal to 80% using a level of significance equal to 5% (for a one-tailed test).

Sideridis et al. 2014 [23] used a series of Monte Carlo simulations to estimate that 50-70 participants would suffice to evaluate functional connectivity in the brain using EF, EEG or fMRI. Sideridis et al.’s simulation involved medium sized models (i.e. 7 latent variables), but the model size is inversely related with sample size. Therefore, larger models, like those in the present study, require fewer participants.

To account for participant drop out during the follow-up period, sample sizes were increased to n=70 children in each group.

### Eligibility criteria

Three groups of parent-child dyads will be recruited: intervention (n=140), control (n=70), and outcome reference (n=70; Table 1).

### Allocation

After recruiting MAM children they will be randomized 1:1 into two groups : Group A and Group B (Table 2) . Computer generated randomized sequences will be used to allocate these children into different groups. The randomized sequences will be generated by a study investigator who will not be directly involved in assigning intervention during enrolling the participants to avoid bias. These sequences will be sealed in white opaque sealed envelopes and will be opened in front of the mother or legal guardian once they consent for the child by the study physician.

**Table 2.**
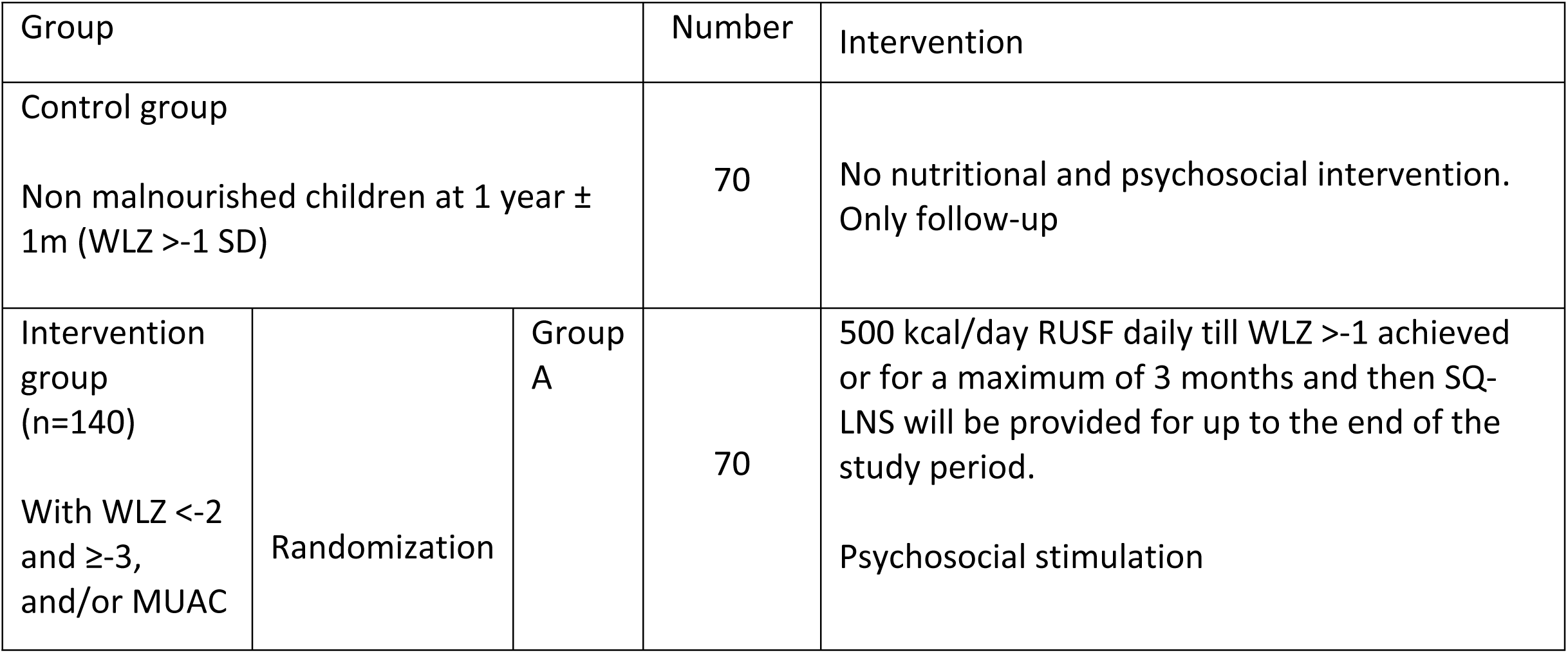
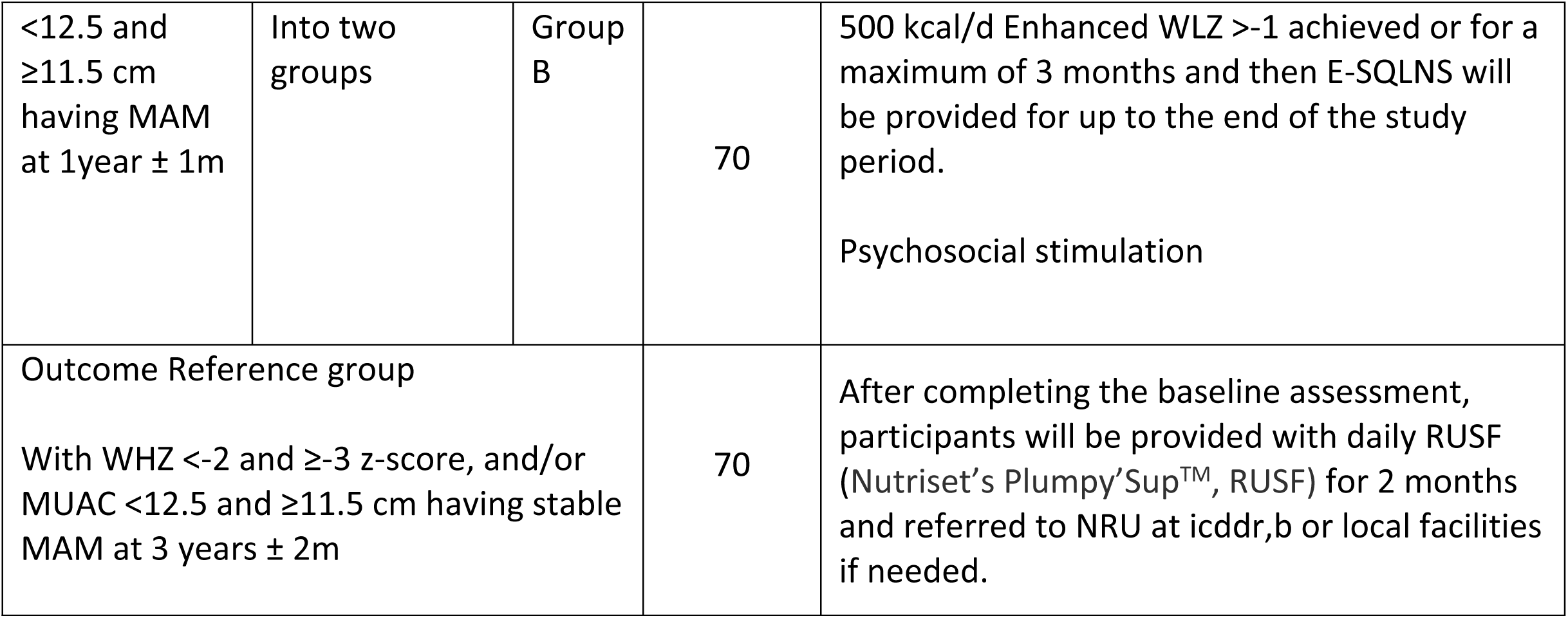
Intervention scheme.

### Intervention

The trial includes a non-malnourished cohort (n=70), MAM intervention cohort (n=140), and MAMs control group (n=70). Children included in the MAMs intervention cohort will be randomized (1:1) to receive either the locally produced Ready to Use Supplementary Food (RUSF) at 500 Kcal/day, or Enhanced Ready to Use Supplementary Food (E-RUSF) at 500 Kcal/day (Table 2). The intervention will be started within one week of completion of baseline assessment.

### Nutritional Supplement

Amongst the several treatment options to manage MAM children, one of the options is to treat these children with RUSF. As per the World Health Organization (WHO), within the Global Nutrition Cluster, discussions centered around a guidance tool known as the “MAM decision tool.” This tool involved exploring various treatment options, one of which included providing Ready-to-Use Food for children aged 6 to 59 months experiencing MAM [24] Several other studies also used RUSF in various clinical trials and found significant improvement in health of MAM children [25]. In Bangladesh, previously rice-lentil based RUSF was locally produced and tested for acceptability [26]. Subsequently, these feeds were utilized in a cluster randomized trial conducted in Bangladesh. The trial involved administering these feeds to 6-month-old children over a year, revealing a significant increase in their average length [27]. The Bangladesh RUSF is an energy-dense rice and lentil-based feed, delivering 250 calories per 50-gram serving, with 45-50% of the calories coming from fat and 8-10% from protein. This RUSF is the Standard-will be produced in a hygienic, sterile food processing laboratory at the Mirpur clinic, by an expert clinical nutritionist, using locally available and culturally acceptable ingredients [26].

E-RUSF (Enhanced-Ready to Use Supplementary Feed adjustment for key nutrients, Nutriset-fr) will be offered to the other 1-year-old intervention group. These feeds contain 24 micronutrients (vitamins and minerals) provided at RDA (Recommended Daily Allowance) levels, functional lipids (Long Chain Polyunsaturated Fatty Acids DHA and EPA), sialylated milk oligosaccharides, neural specific antioxidants (zeaxanthin, lutein; crypto-xanthin) and microbiome modulating dietary soluble fiber mix (inulin + FOS) [28–31].

SQ-LNS, Small Quantity Lipid -based Nutrient Supplements (20gm packets per day) also known as Nutri-butter Plus (Nutriset-fr) will be offered to the children who received the local RUSF for the 2-year follow up period [32]. E-SQLNS (SQLNS enhanced with the same micronutrients in E-RUSF) will be offered to those who received the E-RUSF feed.

All enrolled MAM children will take these feeds daily in addition to their regular complementary feeds at home.

### Dietary Intervention

1. Group A: MAM intervention group participants (n=70) who will be randomized to receive Bangladesh RUSF will receive two packets of that RUSF for consumption providing approximately 500 kcal/day until the child’s weight for length returns to the ‘normal’ range (WLZ >-1), or for a maximum period of 3 months. Once the child’s appetite returns from the high intake drive during rapid catch up to normal intake (which happens when the child reaches their WLZ >-1 ), they will be provided with daily 1 packet SQLNS (20 gm) for the remainder of the 2-year follow-up.
2. Group B: MAM intervention group participants (n=70) who will be randomized to receive E-RUSF will be provided with 1 packet daily providing the same number of calories per day as RUSF, until anthropometric recovery (WLZ > - 1SD) is achieved, or for a maximum of 3 months. Once anthropometric recovery is achieved these individuals will then receive 1 packet of E-SQLNS (26gm) daily until the end of the 2-year follow-up period.

Data from children who do not reach the expected anthropometric recovery (WLZ > - 1SD) after 3 months, will remain in the study as an “Intention to Treat” group. However, they will be referred to the Nutrition Rehabilitation Unit at icddr,b for additional evaluation and management to exclude secondary causes of malnutrition (e.g. Tuberculosis). These children will continue to receive close monitoring with necessary interventions.MAM participants will be followed up for two years to 3 years of age with biological sample collection, anthropometry and neurocognitive assessment at 2 years (22m-26m) and 3 years (34m-38m) of age.

All children will be provided free primary care at the study clinic by designated medical officers and the families will be provided refreshments and will be paid conveyance bills during their visit to study clinic for performing any assessments.

### Psychosocial stimulation

Standard psychosocial stimulation will be provided to the MAM intervention groups. Psychosocial stimulation will consist of bi-weekly, set stimulation curriculum “REACH-UP”. The Reach Up curriculum is a semi-structured, culturally relevant child-age appropriate curriculum that addresses key components of the “nurturing care framework” to promote optimum development [33]. The developmental tasks will take place within a playful environment rather than a work-focused one. During home visits, health-workers will demonstrate to mothers how to play with home-made toys and books and interact with their children aimed at fostering their growth and development.

Toys will be given to the child to play with and learn from, until the next visit, when a new set of developmentally appropriate toys will be provided. The activities in the curriculum are ordered by difficulty level and the health workers will be trained to choose the level for each child according to their ability to do the activities.

### Assessments

Assessments (neuropsychological, behavioral, nutriome, metabolome, and microbiome) will be conducted at baseline, during anthropometric catchup, at 2 and 3 years.

Age, sex, date-of-birth, demography and socio-economic status, medical history, and feeding history will be collected at the time of enrollment.

### Anthropometric data

Weight, length/height, MUAC, head circumference will be measured using established methods (Supplementary materials).

For the intervention group (n=140 children with MAM), anthropometric data will be collected at enrollment, weekly during catch up anthropometric recovery (WLZ >-1), and quarterly for the duration of the study until 36 months of age. Head circumference will be measured at enrollment, at 2 and 3 years of age.

Anthropometric data will also be collected from the control group (n=70 healthy children) at enrollment, then monthly for 3 months, then quarterly for the duration of the study until 36 months of age. Head circumference will be measured at enrollment, at 2 and 3 years of age.

For the 3-year-old MAM control group, anthropometric data will be collected only at enrollment.

### Neuropsychological assessment

Neurodevelopmental assessment data for the control and intervention groups will be collected at enrollment (1year ± 1m), 2years ± 2m, and at 3 years ± 2m. Neurodevelopmental assessment data for the outcome reference group will be collected only at enrollment.

### Executive Functions

Executive functions (EFs) collectively refer to a set of cognitive skills [34, 35] including inhibitory control, planning and cognitive flexibility. Emotion regulation (ER) is an integral part of self-regulation, which is a complex concept that regulates emotions, motivation, cognition (e.g., attention), social interactions, and physical behavior [36].

Our measurements are designed to capture the emergence of executive function at 1 year ± 1m, 2-years ± 2m, and 3-year ± 2m of age. The tests for these age groups are:

- For cognitive flexibility and working memory: Spin-the-Pots, Knock and Tap tasks
- For inhibitory control: Glitter Wand task

### Electroencephalogram (EEG)

The EEG reflects the summation of large populations of neurons that propagate to the scalp surface. These currents can be measured by placing sensors on the scalp [37]. The EEG will be recorded using a NetAmps 300 amplifier system, a 128-channel HydroCel Geodesic Sensor Net (HCGSN, Electrical Geodesics Inc., Eugene, OR) and data acquisition software (NetStation 4.5, EGI). EEG will be used to investigate functional connectivity, power spectrum and event related potentials (ERPs) in both resting-state and during execution of different cognitive tasks.

### Eye tracking

Eye tracking will be used to test distractibility, or disengagement. Children will be presented with an interesting visual stimulus (adapted from Elsabbagh et al) [38] and will be tested on how they disengage their attention from that stimulus. This test has been validated in other low-income settings [39].

A TOBII eye-tracking system (Tobii Studio version 3.3.0) will be used to track participant’s eye movements during cognitive tasks. A small infrared camera (Logitech QuickCam Pro 9000 Webcam V-UBM46 USB Camera Mic Carl Zeiss 2MP) mounted to a computer monitor will track eye movements.

### Functional Near Infrared Spectroscopy (fNIRS)

fNIRS, a non-invasive method suitable for infant studies [40], will be used to measure brain activity that is related to an externally presented stimulus, and to measure activity connections between different parts of the brain with no explicit external stimulus presentation.

fNIRS will be performed using the NTS system (Biomedical Optics Research Laboratory, University College London). This system meets the international standards: IEC 60601 (for medical electrical equipment), and IEC 60825 (for safety of laser products). This system has been used to image brain activity in over 300 healthy infants in London, and 100 infants in Gambia [41].

### Language Environment Analyses (LENA)

The frequency of vocalization or verbalizations and conversational turns in children will be measured. Recording data at multiple time points will provide a measure of the consistency of the language environment. Data will be analyzed using LENA analysis software (LENA Online Version 3.0). The algorithm will determine the number of child and adult vocalizations, conversational turns, and other language measures [42–44].

### Parent Child Interaction (PCI)

A 10-15minute interaction between parent and child will be video recorded and transcribed. Parents will be asked to engage their child in play or in conversations in the presence and absence of toys and books. The child will also play with the toys/books while the parent completes short surveys, or watches a short video. Language transcripts and videos will be coded for the child’s and parent’s use of language and mutual engagement.

### Bayley Scales of Infant and Toddler Development, Fourth Edition (Bayley 4)

Bayley 4 is an extensive formal developmental assessment tool for diagnosing developmental delays in early childhood. Bayley 4 assesses development in children of 1 - 42 months of age in 5 domains: cognition, motor, language, socio-emotional, and adaptive behavior and plays a potential role in assessing EF in infants [45].

### Behavioral assessment questionnaires

SDQ (Strength and Difficulties Questionnaire), PSS (Perceived Stress Scale), FCI (Family Care Indicator), HOME (Home Observation Measurement of the Environment), BRIEF-P (Behavior Rating Inventory of Executive Function - Preschool Version), BISQ (Brief Infant Sleep Questionnaire) – Revised Short Wolke’s behavioral rating and COVID questionnaire and CPAS (childhood psychological adversity questionnaire) will be administered for behavioral assessments of both mother and child at different time points in the study (Supplementary materials). These questionnaires have previously been validated and used in the Bangladeshi population. [46–49]

### Biological samples

Biological samples (blood, stool and buccal swabs) will be collected, prepared and preserved following standard operating procedures (SOPs) created for this protocol (Supplementary materials).

### Blood analytes

Child’s peripheral blood (2-3 ml) will be collected (EDTA vacutainer), separated into plasma and red blood cells and immediately frozen (-80°C), to enable measurement of vitamin and mineral micronutrients, functional lipids, and microbiome derived blood metabolites (Sample code “33”). Peripheral blood samples will be collected from the control and intervention arms: at enrollment 1year ± 1m, 2years ± 2m, and 3 years ± 2m. In the intervention group, an additional sample will be collected during anthropometric recovery or following the 3-month E-RUSF/ RUSF intervention. Peripheral blood samples will be collected from the outcome reference arm group only at enrollment. Samples will be used for low pass whole genome sequencing (Gencove) and discovery metabolomics.

### Stool collection

Child fecal samples will be collected (Zymo Fecal collection tube, OMNImet.GUT ME-200 tube according to the manufacturer’s instructions, sample code “10”). Samples will be used for shotgun metagenomic (20M 250bp reads per sample) sequencing, functional pathway analysis and stool metabolomics. Fecal samples will be collected from the control and intervention arms at enrollment (1year ± 1m, 2years ± 2m, and 3 years ± 2m). Additional samples will be collected during anthropometric recovery for the intervention group and following the 3-month E-RUSF/ RUSF intervention. Stool samples will be collected from the outcome reference arm only at enrollment.

### Buccal Swab

Buccal swabs will be collected using mini-swabs (MS-01) and BuccalFix tubes BFX/S1/05/50 (Isohelix, according to the manufacturer’s instructions). Samples will be coded as “20” for genomic/epigenomic studies. Buccal swabs will be collected from the control and intervention arms at enrollment 1year ± 1m, 2years ± 2m, and 3 years ± 2m. Buccal swab samples will be collected from the outcome reference children only at enrollment.

### Maternal Assessments

1. Weight and height
2. One stool sample (5gm non diarrheal stool sample)
3. One buccal swab sample
4. 5 ml peripheral blood collected in an EDTA tube

These samples will be collected from the mother during the baseline assessments of all enrolled children. SOPs and CRFs for the collection of these maternal biological samples are supplied (Supplementary materials).

### Study risks and confidentiality

Participant confidentiality is rigorously maintained by the involved investigators, their team, the sponsor and their representatives. This confidentiality encompasses records, research data, clinical details of participants and all other data generated during their involvement in the study. Each participant will be assigned a unique study identification number (SID) for use on study documents and in the study database to minimize risks to confidentiality. The document connecting the SID to patients’ names and medical records will be securely stored in a locked office and will not be accessible to individuals not affiliated with the study. All computers containing the study database will be password-protected ensuring that only authorized personnel associated with the study can access the database.

All neuroimaging/behavioral procedures in this study, have been performed in various studies worldwide, are non-invasive, pain free, and are not known to induce adverse effects in participants. Devices have been adapted to the infant population to ensure safety and comfort, and every measure proposed for use in this project is currently being used in the Crypto and PROVIDE Cohorts in Mirpur. Children will be constantly monitored, and the experimenter will pause the session if children become fussy or fidgety. If children become difficult to be placated, the sensors will be removed to end the session.

To minimize the risk of fatigue from the neuroimaging testing, we have split the battery of tests into two sessions, with options for breaks and snacks as required.

### Data monitoring and quality control

A quality control team, comprised of investigators, field research officers and other team members with experience in field activities, will be responsible for regular observations at the study site and checking data for validity and completeness. For 3-5% of study participants, the quality control team will independently repeat data collection, on the same day, using a field-tested format. Errors will be addressed and corrective measures will be undertaken immediately. All data collection forms will be checked, within the evening they were collected, for completeness, accuracy and consistency. Identical forms, definitions and methods will be used throughout the study period. Weekly staff meetings will be held to review the progress of the study.

Behavioral and early learning data will be collected on a selected subset (5% of children) in parallel, by the quality check (Q/C) officer. The Q/C officer will also undertake qualitative scoring and quality check tests on tester performance, using a semi-structured check-list (Supplementary materials). All test information will be reported.

### Adverse events (AE) and Serious Adverse events (SAE)

All AEs and SAEs, related or unrelated to the study feeds, will be reported for this protocol. AE and SAEs will be assessed for severity, relationship to the study intervention, actions taken and outcomes. All SAEs will be reported to the Ethical Review Committee of icddr,b, within 24 hours of the study staff becoming aware of the event. All participants will be provided free primary care for any AEs reported at the study clinic.

### Data Analysis Plan

The study will result in a rich data set consisting of neuropsychological assessments of executive functions and emotion regulation, blood nutriome and metabolome, microbiomes, genetics, nutritional status, age, sex, DOB, demography and SES from family, medical history of these children.

Statistical tests will be conducted to quantify differences in the neurocognitive assessments (e.g. EF, NIRS and EEG responses) between and within the wasted and non-malnourished cohorts, all of whom have varied levels of exposure to adversities (single adversities, composite exposure scores, and income). Three model types will be employed, (a) a t-test for analyzing univariate outcomes, (b) linear mixed models, and (c) structural equation modeling to analyze multivariate outcomes.

T-tests will be conducted with an alpha level of 5% and the distributions of the two samples will be checked for normality, homogeneity of variances and autocorrelation in the residual terms. The False Discovery Rate (FDR) method will be applied to correct for potential multiple comparisons.

Linear mixed effects model will be used to evaluate longitudinal effects with respect to the group differences and the relationship with other earlier biological and/or psychosocial risk factors, particularly focusing on the brain power and functional change in development. Since these measurements are repeated at three time points, mixed models with either random intercept or random slope will be considered.

Structural equation modeling will be used to evaluate multivariate relationships. Model fit will be evaluated using descriptive fit indices such as the Comparative Fit Index (CFI) which work with relatively small sample sizes (Bentler et al,1990) [50].

### Research Ethics Approval

The protocol (registered on ClinicalTrials.gov, study ID number: NCT05629624) was co-developed by researchers from The University of The West Indies, Boston Children’s Hospital, USA, International Centre for Diarrhoeal Disease Research, Bangladesh (icddr,b), and University of Auckland, New Zealand. Ethical approvals were obtained from the Research Review Committee (RRC; August 21, 2021) and Ethical Review Committee (ERC) of icddr,b (protocol no: PR-21084; September 21, 2021), University of Auckland, New Zealand (approval AH23922; for analyses of collected biological samples) and University of West Indies (CREC-MN.51, 21/22).

### Consent

Parents or legal guardians of all participants will provide informed consent after complete disclosure of all study information. Participants can voluntarily withdraw consent at any time point.

### Patient and Public Involvement

Patients were not involved in the design or conduct of this research. Once the trial has been published, efforts will be made to contact and inform participants of the outcomes.

### Data Availability

After the study completes, anonymous post-processed metagenomics sequencing data will be publicly accessible through the Sequence Read Archive on the National Center for Biotechnology Information. Genetic data will be made available through dbgap. De-identified clinical data can be provided for future research upon valid requests submitted to the International Centre for Diarrheal Disease Research, Bangladesh. Requests will have to fall within the original ethics approvals, with appropriate institutional ethics approval. Requestors must consent to a Data Access Agreement, which involves a pledge to solely utilize the data for the designated research proposal and commitments not to make any attempt to identify any individual participants, to securely store and handle the data, and to dispose of the data upon project’s conclusion.

## Discussion

### Strengths and limitations of this study

This study aims to assess the impact of nutritional and psychosocial interventions on the cognitive and emotional well-being of malnourished children in a low-income setting. We are piloting a nutritional intervention to improve brain health and we will use tools that directly measure EF and ER as well as their response to interventions. The study’s findings may be constrained due to the potential influence of psychosocial adversities, unhygienic living conditions, suboptimal sanitation practices, with accompanying infection, and worm infestation as malnutrition often co-exists with these factors.

### Dissemination

Findings from this study will be communicated to the scientific community through publications in peer-reviewed journals and conference presentations. Study participants will be informed of the study findings through the International Centre for Diarrheal Disease Research, Bangladesh.

### Protocol Amendments

Protocol amendments often involve modifications to the study’s procedures, eligibility criteria, data collection methods, or statistical analysis plan. They may also include changes to the informed consent process or adjustments in the number of participants.

It’s crucial to document and communicate protocol amendments transparently to ensure the integrity of the study and to meet regulatory requirements. Institutional Review Boards (IRBs) or Ethics Committees typically review and approve these amendments to ensure that the changes are ethically sound and do not compromise the safety and well-being of study participants. Researchers will keep accurate records of all protocol amendments to maintain the scientific rigor and validity of the study.

## Acknowledgements

The authors would like to acknowledge Professor Micheal Meaney (Singapore Institute for Clinical Sciences, Agency for Science Technology and Research, Singapore), Professor Kirk Erickson (University of Pittsburgh, Pennsylvania, USA), Professor David Cameron-Smith (Victoria university, Melbourne, Australia), Professor Jack Gilbert (University of California San Diego, USA) and Jukka Leppānen (University of Turku, Finland) for discussions and input into the trial design. Icddr,b acknowledges their core donors, Governments of Bangladesh, Canada for providing unrestricted support and commitment to icddr,b’s research effort.

## Author Contribution

TS, TF and JMO co-wrote the manuscript. NR, SK and FT commented on the manuscript. MZ and TF developed the nutritional intervention, JMO, RH, TF, PD, CAN designed the study and analyses and commented on the manuscript.

